# Late presenter thrombolysis for ischemic stroke between 4.5-24 hours after last known well: a retrospective cohort study

**DOI:** 10.1101/2023.06.08.23291176

**Authors:** Lester Y. Leung, Devin Zebelean, Emiliya Melkumova, Katelyn Skeels, Neel Madan, David E. Thaler

## Abstract

**Background:** Two RCTs demonstrated efficacy and safety of IV alteplase for patients with acute ischemic stroke (AIS) who awaken with symptoms or with last known well (LKW) more than 4.5 hours prior to arrival. However, real world experience using CT perfusion (CTP) or DWI-MRI for patient selection in the U.S. is limited. We developed the Tufts Late Presenter Thrombolysis (LPT) protocol to offer alteplase to patients with wakeup stroke, known LKW more than 4.5 hours, or unknown LKW likely more than 4.5 hours and less than 24 hours, using CTP or DWI-MRI to aid patient selection.

**Methods:** We reviewed ED stroke codes from our Comprehensive Stroke Center between 1/1/20-12/31/22 to identify patients treated with alteplase. Data were collected on demographics, comorbidities, LKW-to-treatment time (LTT), imaging modality, imaging findings, NIHSS, vessel occlusions, endovascular therapy (EVT), symptomatic ICH, and 90 days mRS. Outcomes for comparative analyses included process times (door to needle, door to CT) and clinical outcomes (90 day mRS, symptomatic ICH).

**Results:** Forty-three of 118 patients (36%) presenting with AIS and treated with thrombolysis were treated between 4.5-24 hours after LKW. Patients treated in the 4.5 hour window and the later window had similar demographics, comorbidities, NIHSS, and EVT rates. CTP was used in the majority of LPT cases. The median penumbra was 45.04 mL (11.55-83.21), and the median core infarct was 5.92 mL (1.89-16.7). Symptomatic intracranial hemorrhage occurred in one LPT case (2.3%). Favorable mRS (0-1) was achieved by 36% of LPT patients with documented 90 day mRS.

**Conclusions:** A pragmatic protocol offering thrombolysis to late presenters may be safe and achieve favorable outcomes.

## Introduction

While last known well to treatment time remains the major determinant of functional outcome and safety of treatment with acute revascularization therapies in acute ischemic stroke (AIS), the increasingly widespread use of advanced neuroimaging technologies in the hyperacute setting offers the ability to select patients, based on favorable physiology, for thrombolysis and thrombectomy in later time windows. Two randomized clinical trials, WAKEUP and EXTEND, demonstrated safety and efficacy of IV thrombolysis beyond 4.5 hours after last known well (LKW) using DWI-MRI and CT perfusion (CTP), respectively, to evaluate patients who awaken with stroke symptoms or have known LKW beyond 4.5 hours.(1!, 2) A third nonrandomized trial, MR WITNESS, assessed the safety of thrombolysis in patients without a witnessed onset of symptoms likely between 4.5 to 24 hours from LKW using MRI.(3) Nonetheless, the American Heart Association-American Stroke Association guidelines for the early management of AIS only incorporated data from the WAKEUP trial: thus, its provisional recommendations regarding thrombolysis beyond 4.5 hours was based solely on an MRI-informed practice.(4) Accordingly, the safety, efficacy, and logistical considerations of thrombolysis beyond 4.5 hours using CTP in the real world setting is largely unknown. Furthermore, the selection criteria for the WAKEUP and EXTEND trials do not readily overlap, making it challenging to offer thrombolysis in these scenarios while adhering strictly to the RCT selection criteria.

Acknowledging the different target populations, heterogeneous selection criteria, and different imaging modalities used in recent clinical trials on thrombolysis in later time windows, we sought to create a pragmatic, inclusive protocol for “late presenter thrombolysis” at Tufts Medical Center, favoring CTP as the more readily available technology. We started this protocol in February 2020 as a nonmandatory treatment option that our neurologists could offer to select patients. In this study, we sought to compare characteristics of patients treated with thrombolysis through standard and LPT protocols and assess outcomes for patients treated through the LPT protocol.

## Methods

### Design and sample

This is a retrospective cohort study including all patients age ≥ 18 hospitalized at a comprehensive stroke center (Tufts Medical Center) with a principal diagnosis of acute ischemic stroke (AIS) presenting to the emergency department and treated with thrombolysis between 1/1/2020-12/31/2022. An evidence informed protocol (Figure 1) extending the time window for thrombolysis up to 24 hours was implemented on 2/1/20 and allowed for use of CTP or DWI-MRI to aid patient selection. Patients were identified from a prospective database of stroke code activations. The LPT cohort were patients who received alteplase between 4.5-24 hours after LKW, and the standard window comparison group were patients who received alteplase within 4.5 hours after LKW. All patients who survived the hospitalization received phone calls at approximately 90 days post-stroke to assess a modified Rankin Score (mRS). This study was approved by the IRB at Tufts Medical Center and followed STROBE guidelines.

**Figure 1.**
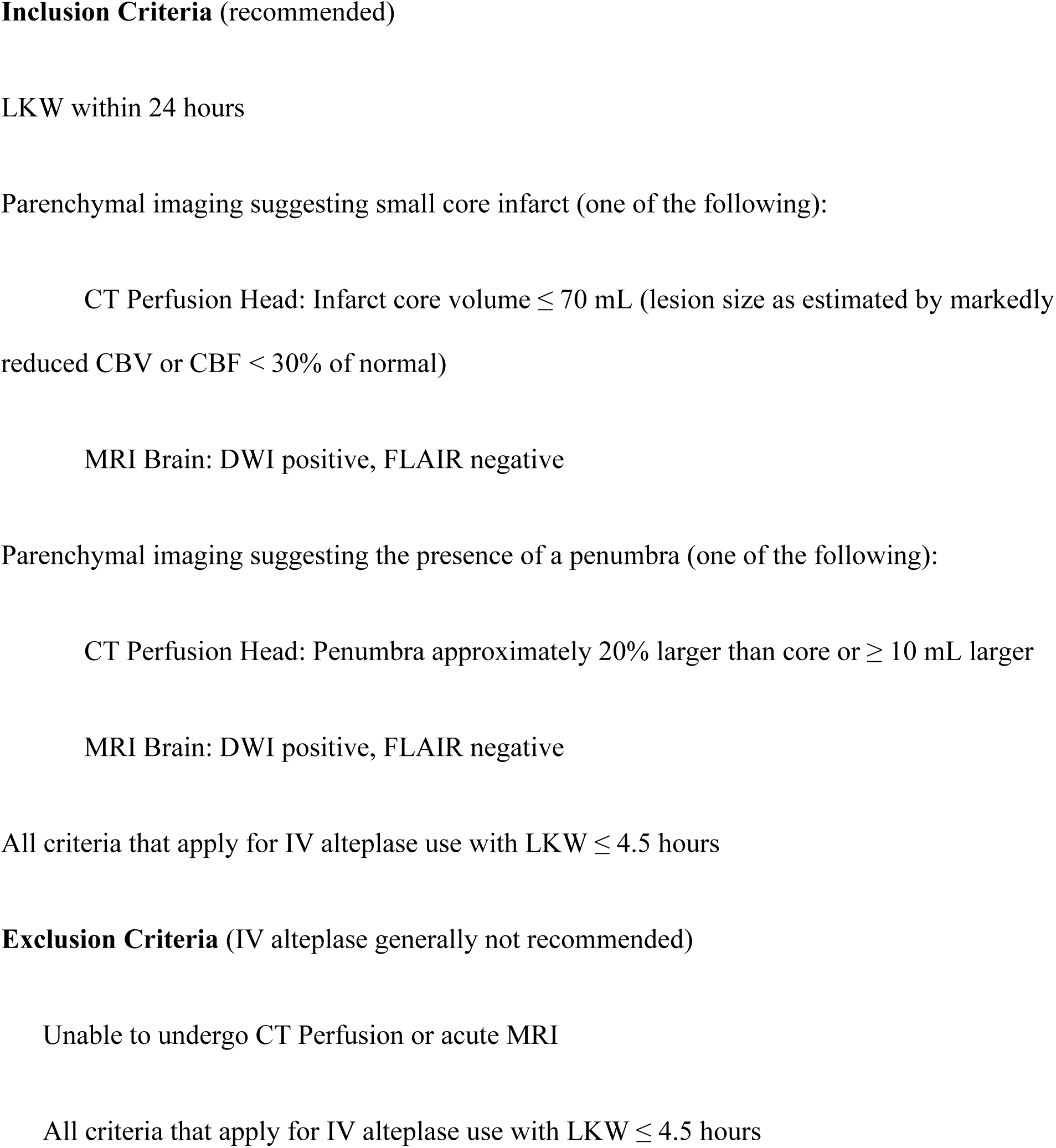
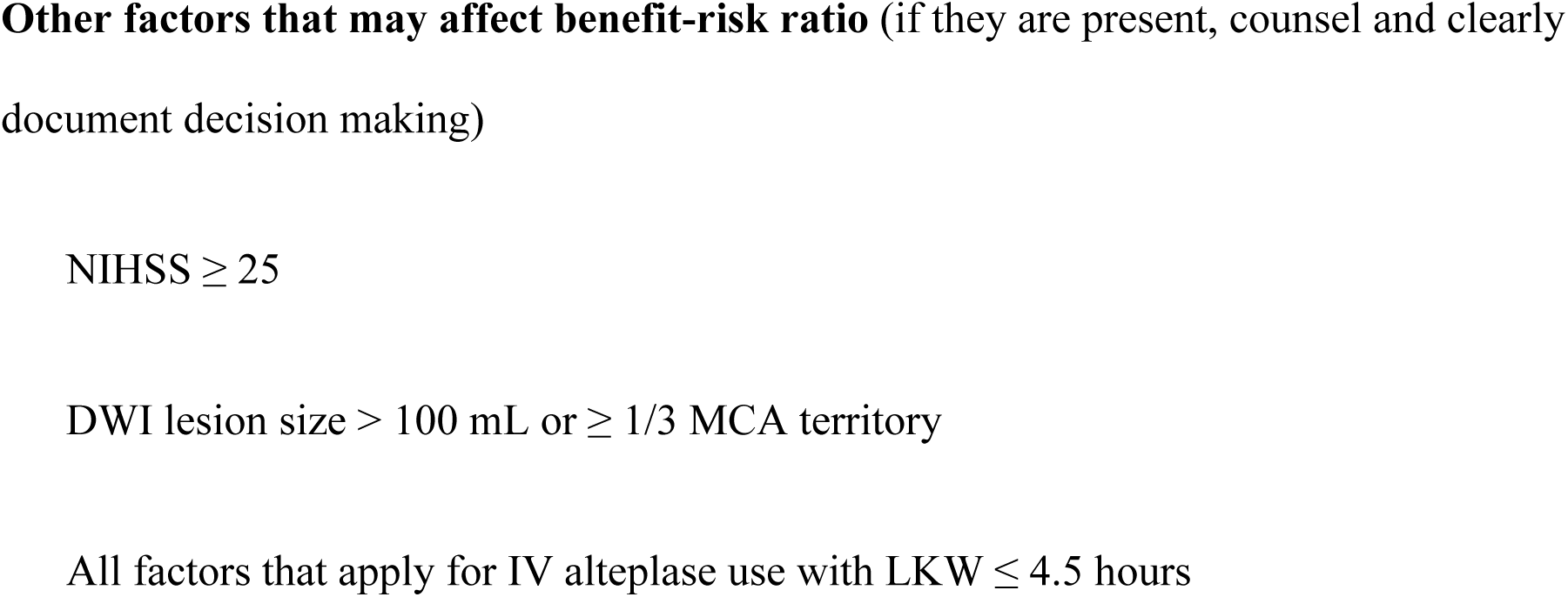
Tufts Late Presenter Thrombolysis Protocol. Thrombolysis for suspected acute ischemic stroke may be offered to patients awakening with stroke symptoms, with LKW > 4.5 hours, and with unknown LKW likely less than 24 hours, with the following selection criteria:

#### Covariates

Data were collected on demographics, comorbidities, admission NIHSS, imaging modalities, imaging findings, LKW-to-thrombolysis time (in hours), endovascular treatment (EVT), symptomatic intracranial hemorrhage (sICH, defined as imaging signs of any intracranial hemorrhage and post-treatment NIHSS increase ≥ 4), and 90 day mRS. CTP imaging was processed using Syngo.via (Siemens) using default settings (core infarct defined as a region of absolute cerebral blood flow [CBF, measured in milliliters of blood per 100 grams of brain] reduction to 30% or less than the maximum CBF; penumbra defined as the region with time-to-maximum [TMax, measured in seconds] greater than 6 seconds). Outcomes for comparative analyses included process times (door to needle, door to CT; measured in minutes) and clinical outcomes (90 day mRS, sICH). Missingness for specific covariates is indicated in the tables, including loss-to-followup for the 90 day mRS assessment.

### Statistical analyses

Descriptive statistics and comparative statistics (t-test, Chi square, Kruskal-Wallis rank sum) are reported according to the structure of the data). R (version 4.2.3, Vienna, Austria) was used for all statistical analyses.

## Results

Among 118 patients presenting to the ED with AIS and treated with alteplase, 43 (36%) were treated with the LPT protocol. Demographics, comorbidities, NIHSS, and EVT rates were similar between both cohorts (Table 1). The median NIHSS was 8 (IQR 3, 14) in both cohorts. The median LKW-to-thrombolysis time was 11 (IQR 8, 15) in the LPT cohort as compared to 2 (IQR 1, 3) in the standard window cohort.

**Table 1.** Baseline characteristics, outcomes, and imaging findings of patients considered for late presenter thrombolysis.

| **Characteristic** | **Standard window**<br>**(**< 4.5 h**)**<br>**(**n=75**)** | **Late presenter**<br>**thrombolysis (wakeup**<br>**stroke, 4.5-24 h)**<br>**(**n=43**)** | **p-value** |
| --- | --- | --- | --- |
| **Age, median in years (IQR)** | 69 (58,80) | 72 (61,80) | 0.69 |
| **Sex, female (%)** | 37 (49.3%) | 18 (41.9%) | 0.43 |
| **Race, non-white (%)** | 21 (28.8%)* | 16 (38.1%)** | 0.30 |
| **Comorbidities** |  |  |  |
| **Atrial fibrillation** | 9 (12.0%) | 8 (18.6%) | 0.33 |
| **Coronary artery**<br>**disease** | 12 (16.0%) | 10 (23.3%) | 0.33 |
| **Diabetes mellitus** | 16 (21.3%) | 15 (34.9%) | 0.11 |
| **Hyperlipidemia** | 41 (54.7%) | 30 (69.8%) | 0.11 |
| **Hypertension** | 54 (72.0%) | 32 (74.4%) | 0.78 |
| **Prior stroke** | 13 (17.3%) | 7 (16.3%) | 0.88 |
| **Tobacco use (ever)** | 18 (24.0%) | 7 (16.3%) | 0.32 |
| **Admission NIHSS, median**<br>**(IQR)** | 8 (3, 14) | 8 (3, 14) | 0.74 |
| **LKW-to-thrombolysis,**<br>**median in hours (IQR)** | 2 (1, 3)** | 11 (8, 15) | <0.001 |
| **Endovascular therapy** | 23 (30.7%) | 8 (18.6%) | 0.15 |
| **performed (%)** |  |  |  |

\*Missing for 2 patients.
\*\*Missing for 1 patient.

Regarding process outcomes, both median door to CT times and median door to needle times longer for patients in the LPT cohort as compared to the standard window cohort (Table 2).

**Table 2.** Clinical and process outcomes for patients treated with standard window and late presenter thrombolysis.

| **Outcome** | **Standard window**<br>**($< 4.5$ h)**<br>**(n=75)** | **Late presenter**<br>**thrombolysis (wakeup**<br>**stroke, 4.5-24 h)**<br>**(n=43)** | **p-value** |
| --- | --- | --- | --- |
| **Door to CT time, median in**<br>**minutes (IQR)** | 22 (17, 29)* | 34 (26, 48) | 0.013 |
| **Door to needle time,**<br>**median in minutes (IQR)** | 42 (32, 60) | 76 (58, 118) | $<0.001$ |
| **Symptomatic ICH (%)** | 0 (0.0%) | 1 (2.3%) | 0.19 |
| **90 day mRS, median (IQR)** | 1 (0, 4)** | 2 (1-3)† | 0.33 |
| **90 day mRS (% 0-1)** | 28 (50.9%)** | 12 (36.4%)† | 0.19 |

\*Missing for 1 patient.
\*\*Missing for 20 patients or 27% (standard window).
†Missing for 10 patients or 23% (LPT).

Clinical outcomes were comparable between both cohorts (Table 2). Favorable 90 day mRS (defined as 0-1) was achieved by 36.4% of patients in the LPT cohort as compared to 50.9% in the standard window cohort (p = 0.19) among those with a documented 90 day assessment. Symptomatic ICH occurred in a single patient in the LPT cohort (2.3%) and in no patients in the standard window cohort (p = 0.19).

Advanced imaging findings are described in Table 3. Among LPT patients, CTP was the advanced neuroimaging modality for 91% of patients. DWI-MRI was used for 1 patient (2.3%), and no advanced imaging was used for 3 patients (7.0%), due to presentation near the 4.5 hour threshold in 2 cases and a very late presentation near the 24 hour mark in 1 case (i.e. CTP was omitted to expedite thrombolysis with a clinical presentation of monocular vision loss due to central retinal artery occlusion). Core infarcts were generally small (median 5.9 mL, IQR 1.9, 17), and penumbras were generally intermediate in volume (median 45 mL, IQR 12, 83). Thrombolysis was offered in 4 cases where CTP did not detect a penumbra or a core infarct (i.e. lacunar stroke) but a clinical deficit was present. Vessel occlusions were present on CT angiography in 65% of cases (with 35% due to large vessel occlusion defined as terminal ICA, M1, or basilar artery occlusion).

**Table 3.** Imaging findings for patients treated with late presenter thrombolysis.

|  |  |
| --- | --- |
| **Advanced imaging performed** | n = 43 |
| **CTP** | 39 (90.7%) |
| **DWI-MRI** | 1 (2.3%) |
| **None** | 3 (7.0%) |
| **Core infarct measurement, median in mL (IQR; range)** | 5.92 (IQR 1.89, 16.7; range 0-83.04) |
| **Penumbra measurement, median in mL (IQR; range)** | 45.04 (IQR 11.55, 83.21; range 0-215.21) |
| **Potential recuperation ratio, median (IQR; range)** | 79.85% (IQR 60.75%, 94.52%; range 0-100%) |
| **Vessel occlusion on CTA** | 28 (65.1%) |
| **ACA (A2)** | 2 |
| **Basilar artery** | 4 |
| **MCA (M1)** | 8 |
| **MCA (M2)** | 5 |
| **MCA (M3)** | 1 |
| **PCA (P2)** | 3 |
| **ICA (terminus)** | 3 |
| **Vertebral (V4)** | 2 |

## Discussion

To our knowledge, this is the first study describing real world experience with a pragmatic protocol leveraging both CTP and DWI-MRI to select a broad range of patients for thrombolysis when presenting between 4.5 and 24 hours after LKW. This builds upon the findings of the WAKEUP and EXTEND trials as well as prior observational studies using noncontrast CT for patient selection.(1-3, 5, 6) Most of the randomized trials informing this practice used MRI as the primary modality to select patients, but this is impractical for many hospitals that lack rapid and consistent access to this technology.(7) This study illustrates how CTP can be used to inform clinical decision-making despite lingering concerns regarding the possible low sensitivity of CTP for detecting small infarcts and posterior circulation infarcts.

The central finding of this study is that thrombolysis beyond the conventional time window can be achieved without a high rate of sICH when patients are selected with advanced neuroimaging guidance. Our pragmatic protocol (e.g. not mandating treatment or strict adherence to specific core or penumbra measurements) allowed for treatment with a variety of clinical and imaging characteristics, including large penumbras, lacunar syndromes, and preceding EVT. Considering the high rate of CTP use versus MRI, this study suggests that a protocol based solely on the WAKEUP trial selection criteria that only allows use of DWI-MRI for patient selection may be too restrictive, especially in hospitals where CTP is more readily available than MRI in a timely fashion. This study adds to the data indicating that thrombolysis beyond 4.5 hours after LKW can be accomplished using CTP and is reasonably safe as compared to standard window thrombolysis.

The process time outcomes in this study illustrate early challenges and opportunities for improvement with the LPT protocol. We anticipated that the door to needle time would be longer for LPT patients as compared to standard window patients due to the need to process and interpret CTP or DWI-MRI findings and incorporate this into treatment decisions. However, the door to CT time was also longer for LPT patients: this time interval ideally should not be different between LPT and standard window patients, so this difference suggests that the Neurology and Emergency Medicine clinicians and Pharmacists might not have proceeded through the stroke code protocols as swiftly for patients potentially eligible for LPT as compared to standard window patients. This may be due to the contemporary novelty of this practice and uncertainty regarding the effectiveness and safety of this practice. The findings from this study hopefully can provide reassurance on the potential for clinical benefit and the relative safety of this practice, and thus encourage stroke teams to proceed through the treatment process as efficiently for LPT patients as with standard window patients given the continued importance of time as a determinant of outcomes in acute stroke.

Notably, the majority of estimated core infarcts for patients treated with the Tufts Late Presenter Thrombolysis protocol were relatively small and comparable to those reported in EXTEND (median 6 mL in our study vs median 5 mL in EXTEND).^2^ Even the one patient who had a sICH had a small estimated core infarct (2 mL with a 72 mL penumbra). However, our protocol (modeled on the EXTEND protocol) used an upper threshold recommendation of 70 mL for estimated core infarcts on CTP. Two patients had estimated core infarcts above the 70 mL recommended threshold (80 and 83 mL), and five additional patients had estimated core infarcts between 20-70 mL; none of these patients had sICH. Altogether, this study provides only limited data on the safety of thrombolysis for patients with medium to large estimated core infarcts on CTP. Future trials of LPT should intentionally assess efficacy and safety of thrombolysis with larger estimated core infarcts than those seen in EXTEND.

This study has important strengths and limitations. First, patients were administered thrombolysis in accordance with a local, standard-of-care protocol informed by prior clinical trial and observational data, so the process of evaluating and treating patients was largely homogeneous. Second, to our knowledge, this is the largest report of thrombolysis cases combining potential treatment-eligible patient categories outside the standard time window.

Third, the diverse spectrum of demographic and clinical characteristics of both cohorts is comparable to many comprehensive stroke centers with broad catchments areas in the U.S. One important limitation is that the Tufts LPT protocol does not mandate thrombolysis in the later time window which could result in some selection bias. Nonetheless, the characteristics of treated patients in both time windows were similar, so any selection bias is unlikely to influence the results of the comparative analyses. Finally, there was considerable missingness in 90 day mRS for patients treated in both time windows, so the proportion of patients achieving excellent outcomes may be subject to loss-to-follow up bias. However, the proportion of missingness was similar between the two cohorts, again suggesting that this is unlikely to substantially influence the comparative results.

## Conclusion

The Tufts Late Presenter Thrombolysis protocol leveraging both CTP and DWI-MRI is pragmatic, feasible, safe, and associated with favorable outcomes for patients with acute ischemic stroke.

## Data Availability

All data from this study is available from the corresponding author upon reasonable request.

## Acknowledgements

We would like to thank Amr Jijakli, MPH, MBBS, and Mohammed Al-Dulaimi, MD for their assistance with data collection.

